# The neurodevelopmental spectrum of *CASK*-related disorder

**DOI:** 10.1101/2024.09.30.24314620

**Authors:** Jessica Martin, Alkistis Mavrogalou-Foti, Josefine Eck, Laura Hattersley, Kate Baker

**Affiliations:** MRC Cognition and Brain Sciences Unit, University of Cambridge; CASK Research Foundation; Department of Medical Genetics, University of Cambridge; Department of Pathology, University of Cambridge

**Keywords:** *CASK*-related disorder, X-linked Intellectual Disability, MICPCH, Epilepsy, Autism characteristics, Adaptive function, Cerebral Visual Impairment, Genetic, Sleep difficulties

## Abstract

**Background:** Pathogenic *CASK* variants are associated with neurodevelopmental disorders of variable severity including X-linked intellectual disability (XLID) and microcephaly with pontocerebellar hypoplasia (MICPCH). Although the number of diagnosed cases is rising, current understanding of the *CASK*-related neurodevelopmental spectrum is limited. Here, we systematically review the published characteristics of individuals with *CASK*-related disorder, and compare these to a more recently-diagnosed group. We provide quantitative information about the ranges of adaptive abilities, motor function, visual function and social-emotional-behavioural characteristics, and explore within-group associations.

**Methods:** 151 individuals with *CASK* variants were identified in published literature. 31 children and young people with *CASK* variants were recruited to the UK-based Brain and Behaviour in Neurodevelopmental disorder of Genetic Origin (BINGO) project. BINGO-participating caregivers completed a bespoke medical history questionnaire and battery of standardised neurodevelopmental measures.

**Results:** Comparing the recently diagnosed BINGO *CASK*-related disorder group to previously reported individuals, we found consistent prevalence of tone abnormalities, sensorineural hearing loss and epilepsy, but lower prevalence of severe/profound ID, MICPCH, optic atrophy and nystagmus. Areas of frequent difficulty not highlighted in previous reports include sleep difficulties and cerebral visual impairment (CVI). Neurodevelopmental characteristics were highly variable within the BINGO *CASK*-related disorder group, and group-wide patterns were similar to those observed in other rare genetic conditions. Within the BINGO *CASK*-related group, epilepsy is significantly associated with ID severity, after controlling for age. Sub-groups with MICPCH or microcephaly only have equivalent ranges of adaptive function, but MICPCH may be associated with more severe motor difficulties.

**Conclusion:** The spectrum of neurodevelopmental characteristics associated with *CASK*-related disorder appears to be broadening with increased access to genome-wide diagnostic testing. Further studies are needed to elucidate the relationships between *CASK* variants, structural brain development, epilepsy, and neurodevelopmental characteristics.

## Background

Calcium/calmodium-dependent serine protein kinase (CASK) is a multi-domain scaffolding protein and a member of the membrane-associated guanylate kinase (MAGUK) family, with diverse neuronal functions (reviewed in 1). Several neurodevelopmental presentations have been reported in association with *CASK* variants including: X-linked intellectual disability (XLID) (2,3), microcephaly with pontine and cerebellar hypoplasia (MICPCH) (4–6), XLID with or without nystagmus and microcephaly (7,8), X-linked optic and ophthalmic problems (9), and FG syndrome (10,11). Prior to next generation sequencing (NGS) diagnostics, most individuals with *CASK*-related diagnoses were identified phenotype-first based on these descriptions. With the application of genotype-first NGS diagnostic approaches and broader ascertainment of individuals with neurodevelopmental presentations for clinical genetic testing, the number of individuals diagnosed with a *CASK*-related disorder is rising. Consequently, the reported *CASK*-associated phenotypic spectrum may not be fully representative. There is an urgent need for an up-to-date phenotypic evidence base which can contribute to variant interpretation and provide accurate information about the nature and range of long-term challenges faced by this patient group.

In particular, there has been limited characterisation of cognitive, adaptive, behavioural and social-emotional functions within *CASK*-related disorder. Almost all reported individuals have developmental delay and intellectual disability (ID) (12), and there have been several reported cases of autism spectrum disorder (ASD) and autism-related traits (13–16). However, no systematic evaluation of neurodevelopmental domains has been published. Standardised measures of motor function, communication and social-emotional functions appropriate for individuals with ID have not been included within any case series.

Additionally, the factors predicting neurodevelopmental variation within *CASK*-related disorder are unclear. Truncating and missense variants have been associated with more severe and less severe presentations respectively, with males being more severely affected than females within either variant group (12). However, there are exceptions to this pattern, such as a nonsense variant identified in a mildly affected male patient (14) and missense variants identified in severely affected females (17,18). Moreover, gender and variant type cannot enable precise predictions of ID severity, because the majority of *CASK*-diagnosed individuals are female and have truncating variants. Beyond genotype, the presence and extent of neuroanatomical abnormalities could be another predictor of neurodevelopmental variation. For example, a relationship has been observed between more severe MRI findings (i.e., MICPCH) and a more severe clinical presentation (i.e., epilepsy, ophthalmologic, motor and speech problems), in males but not females with CASK variants (19,20). Another potential within-group influence on neurodevelopment could be epilepsy, which affects around 50% of individuals with a *CASK* variant (21,22), and can predict developmental abilities in other monogenic conditions (23–26). One preliminary study in *CASK*-associated disorder did not find evidence for a link between epilepsy characteristics and severity of developmental delay (21), but variant type and gender may be confounding factors.

In this paper, we aim to address these gaps and provide a comprehensive phenotypic description of *CASK*-related disorder. First, we present a structured review of published clinical case series, to provide a benchmark for comparison to 31 individuals with *CASK* variants assessed via a structured parent-report questionnaire within a wider research study (BINGO – Brain and Behaviour in Neurodevelopmental disorders of Genetic Origin). This comparison enables us to establish whether the *CASK*-related phenotypic spectrum is changing in parallel with broader ascertainment and genotype-first diagnostics. Second, we present standardised multi-domain neurodevelopmental data for the BINGO *CASK* participant group, to document their range of abilities and difficulties, including relationships between domains. Third, we explore whether neurodevelopmental variation within this group is associated with neuroanatomical abnormalities or epilepsy.

## Methods

### Literature Review

We conducted a review of literature describing the clinical features of individuals with *CASK*-related disorder. Relevant literature published up until May 2024 was identified on Medline (PubMed) and Google Scholar using the search terms “*CASK*”, “*CASK*-related disorders”, “microcephaly”, “MICPCH”, and “pontocerebellar hypoplasia”. Additional papers were identified from bibliographies. We included case series that reported (A) at least three individuals with *CASK*-related disorder who had not been previously described and (B) clinical characteristics assessed via direct evaluation or retrospective review of available medical records. Using these inclusion criteria, we identified 15 papers. We excluded Hayashi et al. (2012)(4) from the review as all individuals were later described in Takanashi et al., (2012)(27). Individuals previously described in Valayannopoulos et al., (2012)(28) and Takanashi et al., (2012)(27) were also excluded from our summary of Burglen et al., (2012)(19) and Hayashi et al., (2017)(5), respectively. Thus, our review includes 14 papers and, to the best of our knowledge, summarises unique individuals with *CASK*-related disorder.

### BINGO data collection

#### Participants

Children and young people (CYPs) with *CASK*-related disorder were recruited to BINGO via an international network of *CASK*-related charities (*CASK* Research Foundation, Angelina *CASK* Neurological Research Foundation, Association Enfants *CASK* France), UK-based charities for rare conditions (Unique, Cambridge Rare Disease Network, Genetic Alliance UK), and regional genetic services in the UK. CYPs were eligible if they were aged at least 3 years old, experienced neurodevelopmental difficulties, and had a pathogenic or likely pathogenic *CASK* gene variant, or a variant of uncertain significance, which was previously fed-back via a clinical genetic services as likely related to clinical presentation. Genetic diagnoses were confirmed via medical records. *CASK* variants were classified as single nucleotide variants (SNVs) or copy number variants (CNVs). SNVs were annotated as either (a) protein-truncating, (b) intronic or (c) missense variants; CNVs were annotated as either (a) intragenic or (b) multi-gene in line with previous literature on pathogenic *CASK* genotypes.

#### Phenotyping methods

Parent-report questionnaires were completed online, by post, or via Zoom or telephone interview according to parent preference. Clinical information was gathered via a study-specific medical history questionnaire (MHQ) covering prenatal, infancy and childhood health, plus acquisition of developmental milestones. Information regarding structural brain abnormalities was obtained via Magnetic Resonance Imaging reports (MRI; *n* = 14), genetic test reports (*n* = 9) and parental reports (*n* = 2). Standardised neurodevelopmental questionnaires and procedures for scoring and analysis are described in Supplementary Table 1.

### Statistical analysis

BINGO data were analysed using R studio, version 2023.3.0.386. Where participants had partial datasets, all available data were included in analyses. Raincloud plots were generated for each questionnaire measure. Cut-off scores were overlaid on each raincloud plot where possible to contextualise the severity of difficulties in *CASK*-related disorder relative to normative populations, or previously reported literature. Within-sample relationships were explored between demographic (age, sex), neurological (epilepsy), developmental (Vinland Adaptive Behaviour Scales (VABS; 29) composite score, Flemish Cerebral Visual Impairment Questionnaire (FCVIQ; 30) total score, and social-emotional-behavioural (total scores on the Social Responsiveness Scale (SRS-2; 31), the Developmental Behaviour Checklist (DBC2; 32), and the Repetitive Behaviour Questionnaire (RBQ; 33)) characteristics using statistical tests appropriate to data type. Non-parametric methods were selected if data violated the assumption of normality, and Kendall’s tau correlation was chosen over Spearman’s rank correlation if ties represented more than 20% of the data. To reduce the likelihood of type I errors, p-values were adjusted for multiple testing by applying a Benjamini-Hochberg False Discovery Rate (B-H FDR) correction. Regression analyses were used to examine the influence of epilepsy on adaptive function (VABS composite score). The assumptions of regression were tested prior to analyses (Supplementary Table 6a and 7a). Descriptive differences were explored between CYPs with different neuroanatomical abnormalities (MICPCH vs. microcephaly only vs. no abnormalities reported). Variables explored were demographic characteristics, VABS composite scores and VABS motor domain scores, FCVIQ total scores and epilepsy prevalence.

## Results

### Review of published cases

Supplementary Table 2 summarises the clinical characteristics of 151 individuals carrying *CASK* variants, reported in previous publications. The majority were female (72.2%) and age ranged from 2 weeks to 49 years old at the time of reporting. SNVs accounted for 71.5% of *CASK* variants, encompassing protein-truncating (44.4%), intronic (35.2%), and missense (20.4%) variants. CNVs (28.5%) included both intragenic (48.8%) and multi-gene (51.2%) deletions of varying sizes. De novo variants (77.3%) were more common than inherited variants (22.7%).

The most frequently reported clinical problems were feeding difficulties (57.1%), including gastroesophageal reflux, sucking and swallowing difficulties. Interventions such as thickened liquids/pureed foods, nasogastric tube or gastrostomy were common, although feeding difficulties were noted to improve with age in some individuals (1,20). Tone abnormalities were also frequent, including hypotonia (57.1%), hypertonia (41.8%) or mixed tone abnormalities (14%). Epilepsy was reported in 48.9% overall. Seizure details were inconsistently reported, but a range of seizure types were described, including early infantile epileptic encephalopathies with variable EEG phenotypes (e.g., West Syndrome, Ohtahara syndrome), infantile spasms, later-onset epilepsies (generalised or focal) and myoclonic epilepsies. Many individuals had mixed seizure types and there was a variable response to anti-seizure medications from remission to intractable. Optic atrophy (28.6%), strabismus (30.4%) and nystagmus (25%) were common ophthalmological abnormalities co-occurring with each other. Other reported vision-related phenotypes included astigmatism, myopia, hyperopia, retinopathy, glaucoma and cerebral visual impairment (CVI). Sensorineural hearing loss was present in 24.6%. Other characteristics less frequently reported were scoliosis, kidney problems and sleep disturbances.

The majority of reported individuals with *CASK*-related disorder had severe or profound developmental delay (88%). 56.6% were able to sit without support and acquired this ability between 6 months and 3 years. Fewer individuals (25.7%) had acquired the ability to walk independently with age of acquisition ranging from 18 months to 6 years. There were also three reports of regression of motor abilities, including a case of mild regression at 2 years (6), an individual that lost ability to walk at 25 years after severe febrile seizures (1), and an individual that lost ability to sit after scoliosis surgery (14). A minority of individuals were reported to use some vocal communication (33.8%), which ranged from babbling and vocalisations to short sentences. Two papers reported individuals using Augmentative and Alternative Communication (AAC) systems as non-verbal methods of communication, such as an eye gaze device and Picture Exchange Communication System (PECS; 14,19).

In the papers we identified, there was a limited focus on social, emotional and behavioural problems with only four papers describing social or emotional-behavioural difficulties, such as ASD (1,14), self-injurious behaviours (19) and general behavioural difficulties (28).

Where MRI data were reported, the majority of individuals with *CASK* variants had MICPCH (87.1%). There were also five individuals with cerebellar and/or pontine hypoplasia only (3.6%), four individuals with microcephaly and other brain abnormalities (2.9%), three cases of microcephaly with cerebellar hypoplasia only (2.1%), three of microcephaly only (2.1%), and three reports of no brain abnormalities (2.1%).

### Comparison between reviewed cases and BINGO *CASK*-related group

Thirty-one CYPs with *CASK*-related disorder participated in BINGO and were included in this analysis. Comparison between demographic and clinical characteristics of BINGO participants and the reviewed population is summarised in Table 1. A higher proportion of the BINGO group were female, and the age range was narrower due to an age limit for BINGO eligibility. The proportion of individuals with either *CASK* SNVs or CNVs were near identical between reviewed cases and BINGO, although the BINGO group contained a higher proportion of truncating SNVs versus missense SNVs, and higher proportion of intragenic versus multi-gene CNVs. There were fewer reports of MICPCH in the BINGO group (44.0% vs. 87.1%) and more reports of microcephaly only (32.0% vs. 2.1%).

**Table 1.**
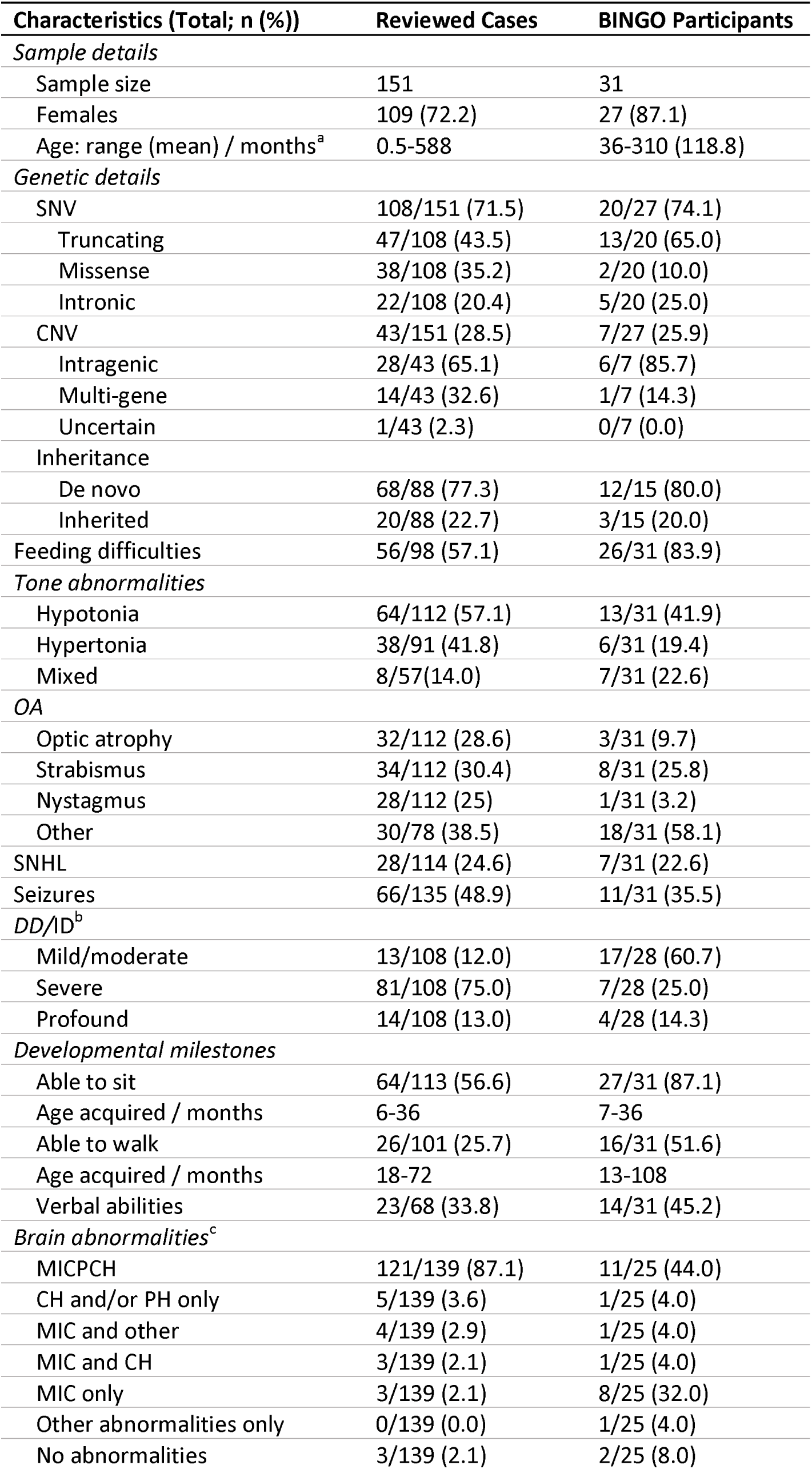
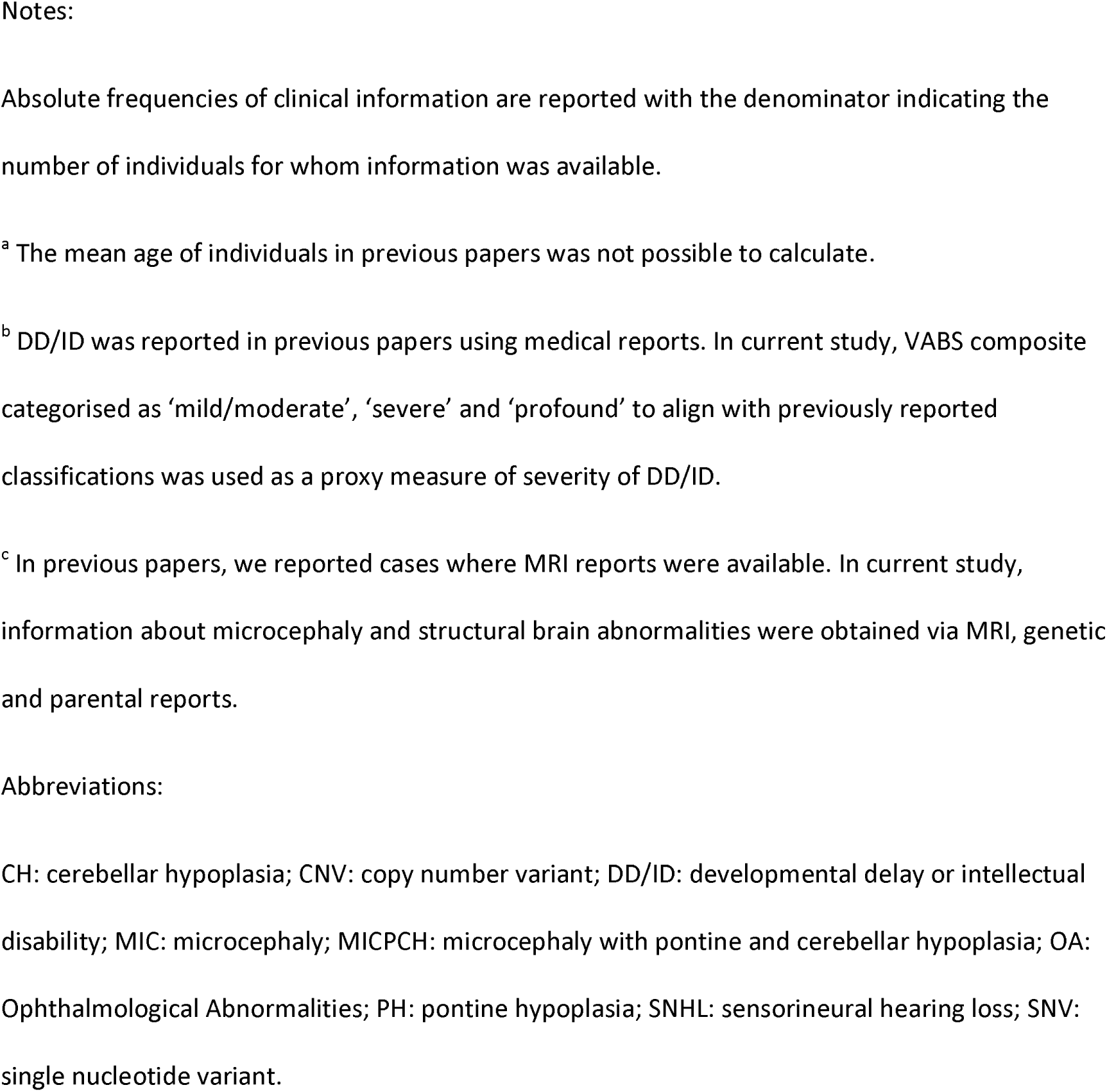
Comparison between reviewed cases and BINGO *CASK*-related disorder group.

| **Characteristics (Total; n (%))** | **Reviewed Cases** | **BINGO Participants** |
| --- | --- | --- |
| *Sample details* |  |  |
| Sample size | 151 | 31 |
| Females | 109 (72.2) | 27 (87.1) |
| Age: range (mean) / months<sup>a</sup> | 0.5-588 | 36-310 (118.8) |
| *Genetic details* |  |  |
| SNV | 108/151 (71.5) | 20/27 (74.1) |
| Truncating | 47/108 (43.5) | 13/20 (65.0) |
| Missense | 38/108 (35.2) | 2/20 (10.0) |
| Intronic | 22/108 (20.4) | 5/20 (25.0) |
| CNV | 43/151 (28.5) | 7/27 (25.9) |
| Intragenic | 28/43 (65.1) | 6/7 (85.7) |
| Multi-gene | 14/43 (32.6) | 1/7 (14.3) |
| Uncertain | 1/43 (2.3) | 0/7 (0.0) |
| Inheritance |  |  |
| De novo | 68/88 (77.3) | 12/15 (80.0) |
| Inherited | 20/88 (22.7) | 3/15 (20.0) |
| Feeding difficulties | 56/98 (57.1) | 26/31 (83.9) |
| *Tone abnormalities* |  |  |
| Hypotonia | 64/112 (57.1) | 13/31 (41.9) |
| Hypertonia | 38/91 (41.8) | 6/31 (19.4) |
| Mixed | 8/57 (14.0) | 7/31 (22.6) |
| *OA* |  |  |
| Optic atrophy | 32/112 (28.6) | 3/31 (9.7) |
| Strabismus | 34/112 (30.4) | 8/31 (25.8) |
| Nystagmus | 28/112 (25) | 1/31 (3.2) |
| Other | 30/78 (38.5) | 18/31 (58.1) |
| SNHL | 28/114 (24.6) | 7/31 (22.6) |
| Seizures | 66/135 (48.9) | 11/31 (35.5) |
| *DD/ID<sup>b</sup>* |  |  |
| Mild/moderate | 13/108 (12.0) | 17/28 (60.7) |
| Severe | 81/108 (75.0) | 7/28 (25.0) |
| Profound | 14/108 (13.0) | 4/28 (14.3) |
| *Developmental milestones* |  |  |
| Able to sit | 64/113 (56.6) | 27/31 (87.1) |
| Age acquired / months | 6-36 | 7-36 |
| Able to walk | 26/101 (25.7) | 16/31 (51.6) |
| Age acquired / months | 18-72 | 13-108 |
| Verbal abilities | 23/68 (33.8) | 14/31 (45.2) |
| *Brain abnormalities<sup>c</sup>* |  |  |
| MICPCH | 121/139 (87.1) | 11/25 (44.0) |
| CH and/or PH only | 5/139 (3.6) | 1/25 (4.0) |
| MIC and other | 4/139 (2.9) | 1/25 (4.0) |
| MIC and CH | 3/139 (2.1) | 1/25 (4.0) |
| MIC only | 3/139 (2.1) | 8/25 (32.0) |
| Other abnormalities only | 0/139 (0.0) | 1/25 (4.0) |
| No abnormalities | 3/139 (2.1) | 2/25 (8.0) |

Notes:
Absolute frequencies of clinical information are reported with the denominator indicating the number of individuals for whom information was available.
<sup>a</sup> The mean age of individuals in previous papers was not possible to calculate.
<sup>b</sup> DD/ID was reported in previous papers using medical reports. In current study, VABS composite categorised as 'mild/moderate', 'severe' and 'profound' to align with previously reported classifications was used as a proxy measure of severity of DD/ID.
<sup>c</sup> In previous papers, we reported cases where MRI reports were available. In current study, information about microcephaly and structural brain abnormalities were obtained via MRI, genetic and parental reports.
CH: cerebellar hypoplasia; CNV: copy number variant; DD/ID: developmental delay or intellectual disability; MIC: microcephaly; MICPCH: microcephaly with pontine and cerebellar hypoplasia; OA: Ophthalmological Abnormalities; PH: pontine hypoplasia; SNHL: sensorineural hearing loss; SNV: single nucleotide variant.

A higher percentage of feeding difficulties was reported in BINGO (83.9% vs. 57.1%), but the range of difficulties and interventions reported were similar to the reviewed group. The frequency of reported tone abnormalities, sensorineural hearing loss and epilepsy were comparable between the BINGO group and reviewed group. A range of seizure types were reported in BINGO, including infantile spasms, myoclonic epilepsies, as well as focal, complex and absence seizures. The age of seizure onset varied from 4 months to 10 years. Reports of strabismus, refractive errors and CVI were similar between BINGO and reviewed cases, but optic atrophy and nystagmus were not common in the BINGO group (9.7% and 3.2%, respectively) compared to reviewed cases (28.6% and 25%, respectively). Sleep difficulties were common in our sample (74.12% ever experienced) but were not widely reported in previous literature (see 19 for exception). Parents reported a range of sleep-associated problems including difficulty settling, regular waking, and need for more or less sleep than expected for age, as well as sleep apnoea.

There were notable differences between BINGO and the reviewed group in acquisition of developmental milestones, whereby a higher percentage of the BINGO group had acquired the ability to sit (87.1% vs. 56.6%) and walk (51.6% vs. 25.7%). Reported verbal abilities were similar across the two groups. Categorisation of ID severity also differed between groups, with a higher percentage of the BINGO group in the mild or moderate (60.7% vs. 12.0%) rather than severe (25.0% vs. 75.0%) category, noting that this categorisation was based on VABS data in BINGO but clinical report in the reviewed sample.

### Neurodevelopmental variation within the BINGO *CASK*-related group

Figures 1-5 show the distribution of scores on neurodevelopmental and social-emotional-behavioural measures. VABS composite scores indicated that global adaptive ability ranged from mildly impaired to severely/profoundly impaired (Figure 1). Communication was the most affected subdomain, whereas Socialisation was a relative strength.

**Figure 1.**
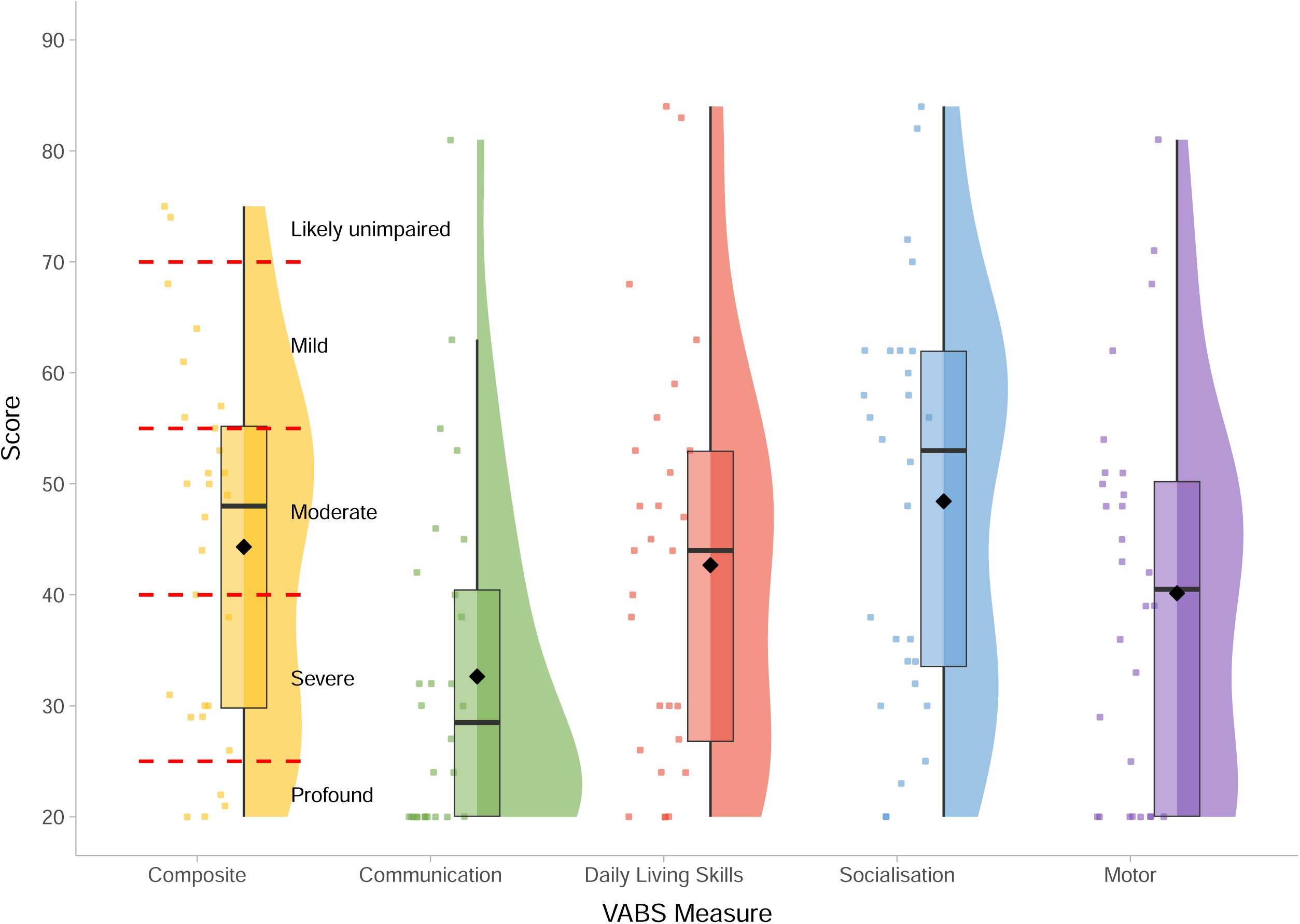
Vineland Adaptive Behaviour Scales - age standardised scores within the BINGO *CASK*-related disorder group. *Legend:* ID severity cut-offs are overlaid in red and annotated in black for the VABS composite score. Mean scores on each measure are represented by the black diamond.

Scores on the SRS-2 revealed high levels of autism-related traits, varying in severity within the atypical range. CYPs with *CASK*-related disorder demonstrate relative strengths in social motivation on the SRS-2, compared to other social subscales or Restricted and Repetitive Behaviours (Figure 2). These patterns are overall similar to observations in other rare genetic conditions (34,35). The majority of DBC2 scores were in the moderate-serious concerns range (Figure 3), with relatively greater difficulties with self-absorbed behaviour (a DBC-2 subscale involving diverse items including mouthing objects, special interests and stereotyped behaviours). On the RBQ, the majority of participants scored above the threshold for *‘*normal’ levels of repetitive behaviours, and scored relatively higher on the stereotyped subdomain (Figure 4). Challenging Behaviour Questionnaire (CBQ, 36) reported rates of self-injury (42.86%), physical aggression (46.43%) and stereotyped behaviour (50.00%) were high (Supplementary Table 3), but similar to rates observed in other rare neurogenetic conditions (37,38). The severity of these behaviours also varied, but on average was similar to those reported in individuals with other rare neurogenetic conditions (39).

**Figure 2.**
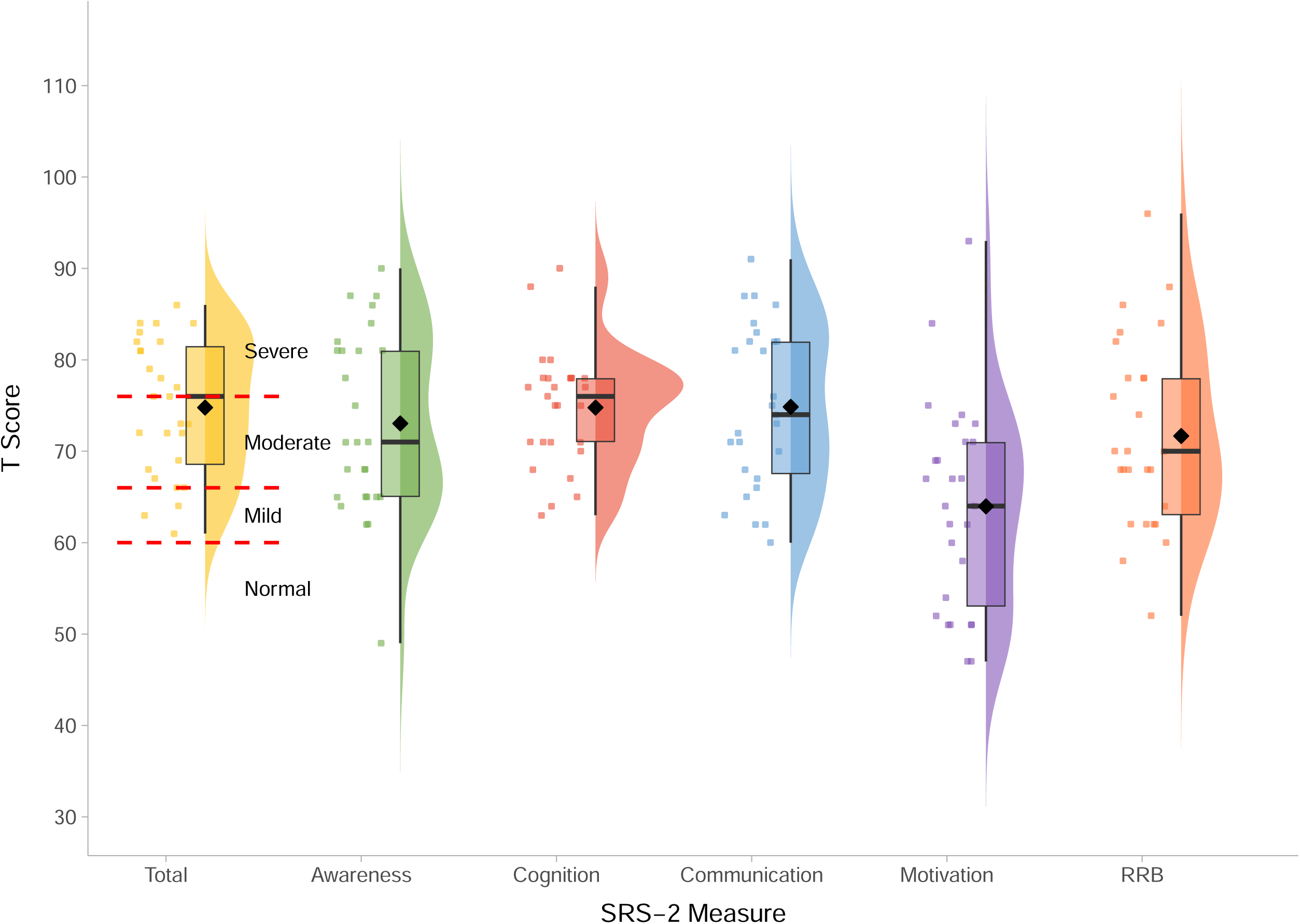
Social Responsiveness Scale - T-scores within the BINGO CASK-related disorder group. *Legend:* Severity cut-offs are overlaid in red and annotated in black for the SRS-2 total score. Mean scores on each measure are represented by the black diamond.

**Figure 3.**
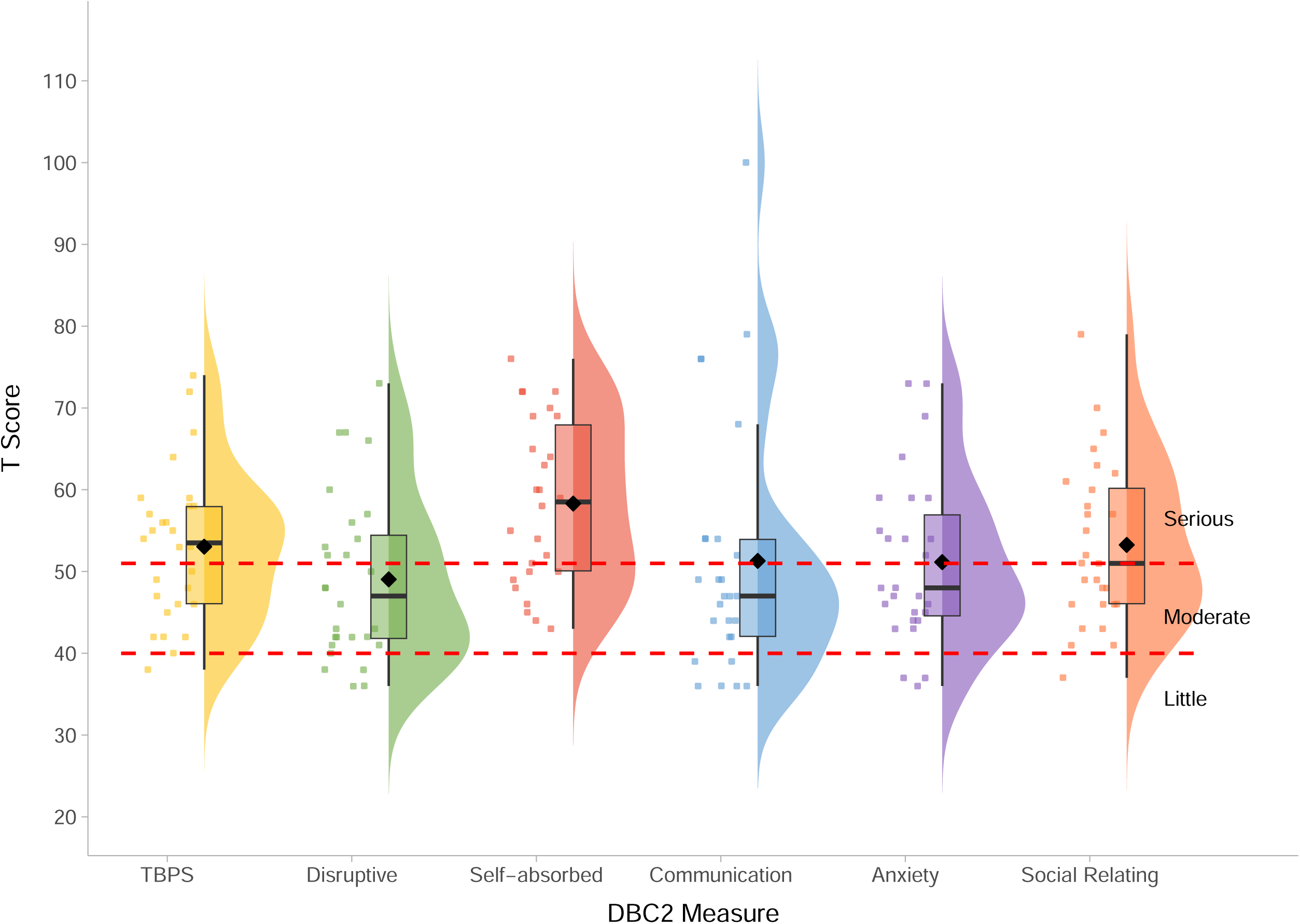
Developmental Behaviour Checklist - T-scores within the BINGO *CASK*-related disorder group. . *Legend:* Levels of concern are overlaid in red and annotated in black for all DBC2 measures. Mean scores on each measure are represented by the black diamond.

**Figure 4.**
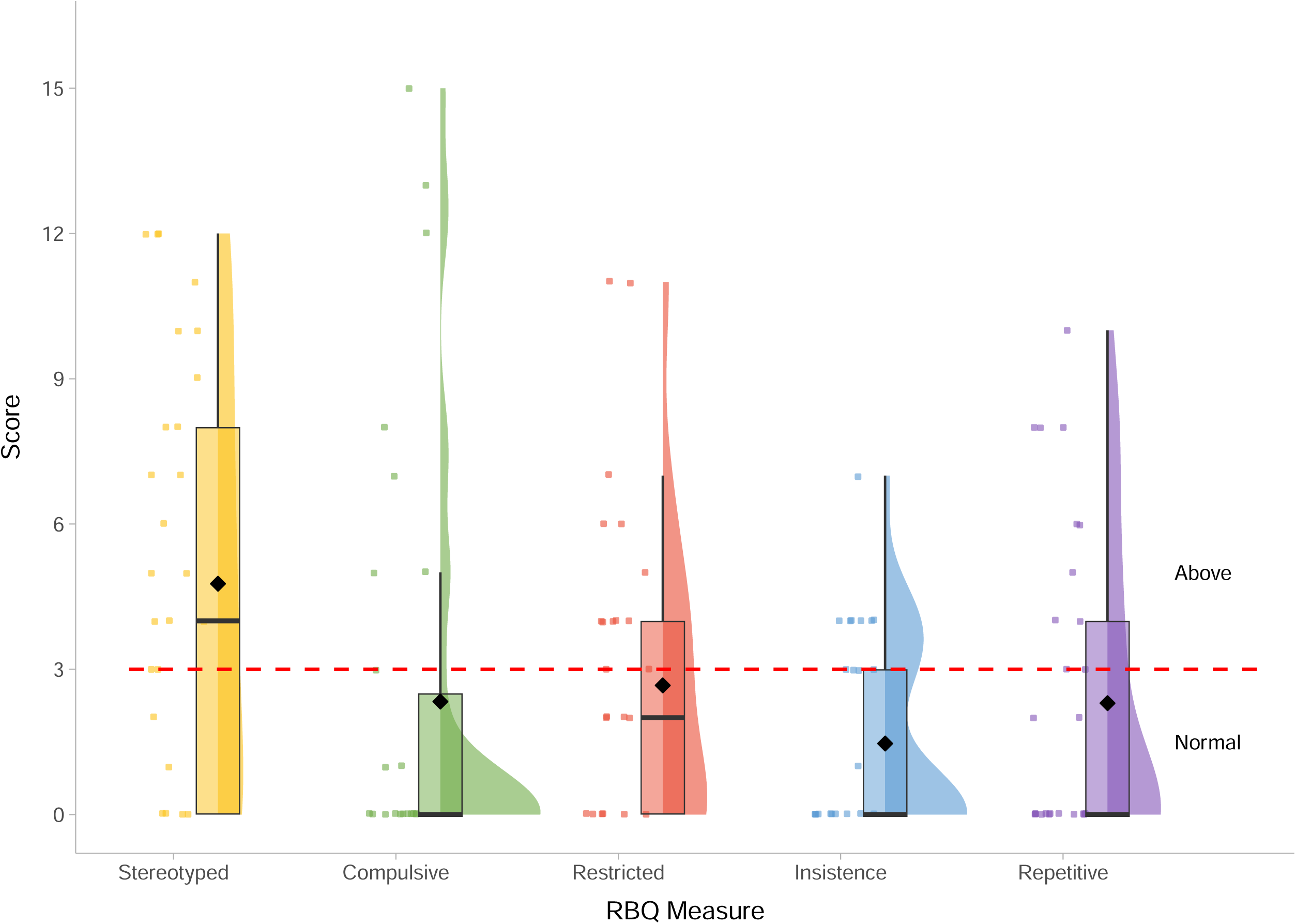
Restricted and repetitive Behaviour Questionnaire – Scores within the BINGO *CASK*-related disorder group. *Legend:* Severity cut-offs are overlaid in red and annotated in black for all RBQ measures. Mean scores on each measure are represented by the black diamond.

**Figure 5.**
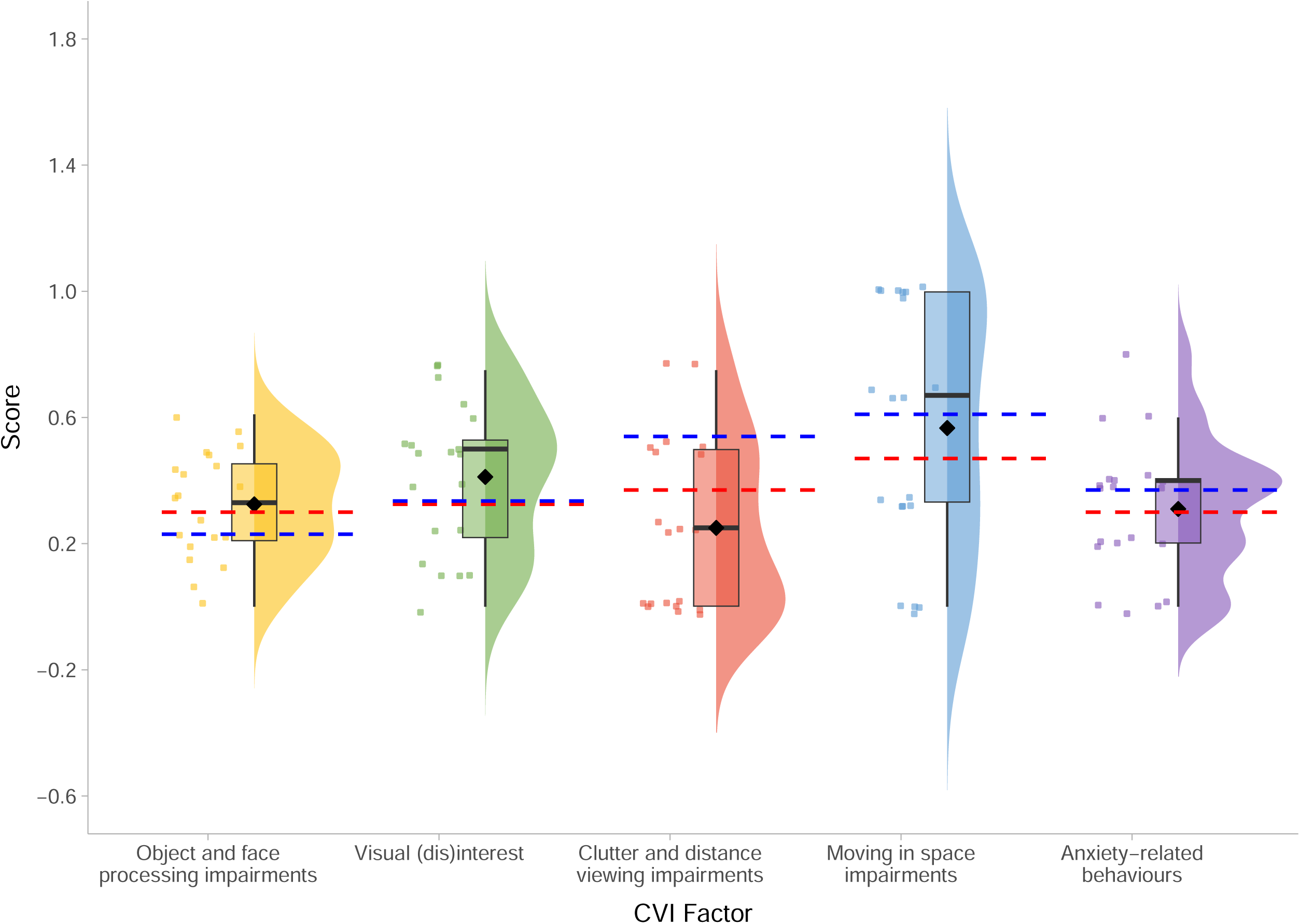
Flemish cerebral visual impairment questionnaire – Factors scores within the BINGO CASK-related disorder group. *Legend:* Mean scores for children with CVI (taken from 32) are overlaid in red, and mean scores for children with CVI and unilateral (taken from 33) are overlaid in blue. Mean scores on each measure for the BINGO *CASK*-related group are represented by the black diamond.

The majority of CYPs with *CASK*-related disorder are at risk of CVI, according to FCVIQ Sum Scores (*M* = 5.10, *SD* = 1.21, *range*: 2-6; 90% above cut-off score of 4). Factor scores on the FCVIQ show that the BINGO *CASK*-related group experienced difficulties with object and face processing, visual interest, moving in space and anxiety-related behaviours of a level comparable to children with CVI, whereas clutter and distance viewing might be relatively spared (Figure 5). Broadening to other aspects of sensory development, most participants scored within the *‘like the majority of others’* range on the Seeking and Avoiding subdomains of the Short Sensory Profile (SSP-2; 40) (Supplementary Table 4), but within the *‘more than others’* and the *‘much more than others’* range on the Sensitivity and Registration subdomains, respectively. The majority of participants also scored within the *‘more than others’* and the *‘much more than others’* ranges on the sensory and behavioural section of the SSP-2, indicating that CYPs with *CASK*-related disorder display more sensory-related behaviours than most children.

### Associations within the BINGO *CASK*-related group

Figure 6 and Supplemental Table 5 show the statistical relationships between demographic, neurological, developmental and social-emotional-behavioural characteristics within *CASK*-related disorder. We found moderate positive correlations between reversed VABS composite scores and (1) age (*r*_s_ = 0.78, *p.adj* < .01), and (2) epilepsy (*r*_pb_ = 0.55, *p.adj* = .03). We also found a moderate positive correlation between total scores on the DBC2 and RBQ (τ = 0.55, *p.adj* < .01). No other relationship survived the B-H FDR correction for multiple testing.

**Figure 6.**
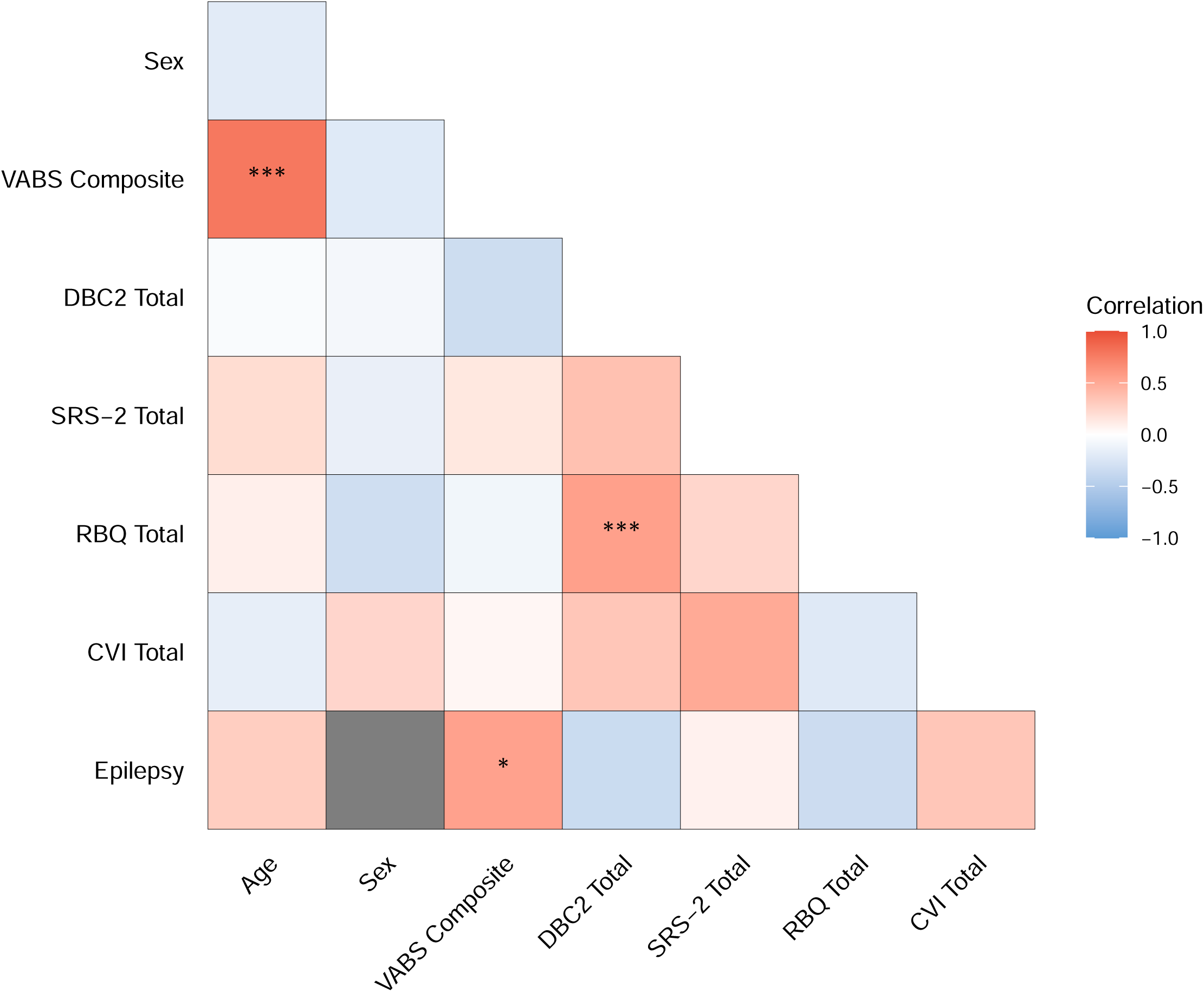
Correlations between demographic and neurodevelopmental domains within *CASK*-related disorder.

When exploring factors that might contribute to neurodevelopmental variation within *CASK*-related disorder, we found epilepsy was a significant predictor of adaptive ability (*F*(1, 25) = 10.69, p < .001), and explained approximately 27% of the variance in adaptive ability (Supplementary Table 6b). We also examined whether entering age as a predictor into this model would affect the relationship between epilepsy and adaptive ability, as we found age was significantly positively correlated with adaptive ability. The overall model was significant (*F*(2, 24) = 16.38, *p* < .001) and explained approximately 54% of the variance in adaptive ability. As shown in Supplementary Table 7b, epilepsy remained a significant independent predictor of adaptive ability (β = 11.00, *p* = .032) and age was also a significant independent predictor of adaptive ability (β = 1.56, *p* < .001).

In addition to epilepsy, available data facilitated descriptive exploration of whether neuroanatomical abnormalities might contribute to neurodevelopmental variation within *CASK*-related disorder. For the purposes of this analysis, we combined MRI findings into three categories: *MICPCH* (i.e., individuals with reported microcephaly and pontocerebellar abnormalities), *Microcephaly* (i.e., individuals with reported microcephaly but without reported pontocerebellar abnormalities), and *Normal* (i.e., individuals without reported microcephaly or neuroanatomical abnormalities). Individuals for whom no brain abnormality data were available, were classified as *NA*. Groups were not large-enough for statistical analysis, however, as shown in Supplementary Table 8, average scores on the VABS composite were similar between individuals with MICPCH (*M* = 40.15) and microcephaly only (*M* = 43.62), but average scores on the motor subdomain of the VABS differed, whereby individuals with MICPCH scored lower (*M* = 30.54) than individuals with microcephaly only (*M* = 46.88). Parent-reported epilepsy was also more common in individuals with MICPCH (53.85%) than microcephaly only (33.33%). Only two individuals were documented as having no neuroanatomical abnormalities, hence comparison to this group is not warranted.

## Discussion

Previous studies of *CASK*-related disorder are largely based on retrospective medical reports of individuals identified phenotype-first, with few involving real-time assessments of developmental ability or social-emotional-behavioural symptoms. To overcome these knowledge gaps for the current and future population of individuals diagnosed with *CASK*-related disorder, we first carried out a comprehensive review of the published clinical characteristics and compared prevalence rates to a more recently diagnosed group of CYPs with *CASK* variants (> 60% diagnosed after 2015). We then evaluated the range of developmental abilities and social-emotional-behavioural problems within this group, using standardised measures validated for individuals with ID. Lastly, we explored potential predictors of neurodevelopmental variation within *CASK*-related disorder, to identify possible markers of prognosis, or mediators of developmental trajectory.

Comparison of the BINGO *CASK*-related group to previous literature provides preliminary evidence for an expanding *CASK*-related phenotype. A lower frequency of developmental delay, severe ID, and characteristics previously considered typical for *CASK*-related disorder (e.g., MICPCH, optic atrophy, nystagmus) were observed in the BINGO group, suggesting the within-group distribution of neurodevelopmental impairments has shifted to include individuals with less severe presentations. These differences could be attributed to sample ascertainment and measurement. CYPs in BINGO were recruited genotype-first, with broad inclusion criteria, reflecting identification of patients in clinical settings i.e. genetic testing for a wide range of developmental concerns, rather than discreet neuroanatomical or ophthalmological diagnoses. Assessment of ID severity varies between medical centres and countries due to differing guidelines and practice. Similarly, accessibility of neuroimaging and reporting of neuroradiological assessments is inconsistent. Adoption of standardised measures, like the VABS in this study, or routine assessment of neuroanatomy could help progress our understanding of *CASK*-related characteristics across cohorts and time. Nevertheless, the clinical spectrum of *CASK*-related disorder within BINGO may more closely reflect the currently-diagnosed *CASK*-related disorder population.

We also identified characteristics associated with *CASK*-related disorder not highlighted in previous case series, which may require increased clinical attention. For example, sleeping difficulties, such as regular night waking and difficulty settling, were commonly reported in our sample of CYPs with *CASK*-related disorder, but were not described in previous literature (with the exception of 19). Specific sleep disorders, along with poor sleep quality, timing, and duration, are common in individuals with ID and rare genetic syndromes (41–43), and parents of individuals with rare genetic conditions frequently highlight sleep as a significant area of concern (44,45). Moreover, poor sleep can be associated with poor adaptive abilities (46,47) and increased challenging behaviour (48–50). As such, it is possible that poor sleep might contribute to observed adaptive and social-emotional-behavioural difficulties in CYPs with *CASK*-related disorder. However, the assessment of sleep difficulties in this study was limited and future studies should strive to generate objective descriptions of sleep difficulties and specific sleep disorders, as well as factors contributing to sleep disturbance such as pain or epilepsy, which are imperative for effective management.

We also found that the majority of CYPs with *CASK*-related disorder were at risk of CVI (according to FCIVQ Sum Scores), however clinical assessments are needed to confirm these findings as only 5 participants in this sample had parent-reported CVI. CVI has been commonly reported in other monogenic conditions with similar genotypic or phenotypic characteristics (51,52). There is a strong association between CVI and seizures across monogenic conditions, in particular infantile spasms (52). This may be particularly relevant for *CASK*, as infantile spasms are the most common seizure type within the group (21).

Despite high levels of difficulties across adaptive and social-emotional-behavioural measures, few significant relationships between measures were identified in the BINGO group of CYPs with *CASK*-related disorder. In other words, difficulties in one developmental dimension do not necessarily go alongside difficulties in another dimension, and categorisation of individuals with *C A S*-*K*related disorder as being mildly or severely affected overall may be overly simplistic. A lack of association could suggest that different characteristics, for example autism-related traits and CVI, might be unique dimensions of neurodevelopmental variability in *CASK*-related disorder. However, we only examined the relationships between total scores on these measures and it is possible that subdomains of autism-related traits and CVI, or specific adaptive functions and challenging behaviours, might be related. Further analysis of the relationships between dimensions using additional methods, in a larger *CASK*-related group and over time, are necessary to understand potential developmental links.

Prognostic markers of developmental ability are of interest to parents and clinicians in order to plan support needs and consider potential interventions. We found that the presence of epilepsy predicts poorer adaptive ability in *CASK*-related disorder. One possible interpretation is that seizure activity negatively impacts cognitive development in *CASK*-related disorder, a relationship also observed in other rare genetic conditions (23–26). Alternatively, epilepsy and poorer adaptive ability might both reflect altered trajectories of brain development, but are not causally related to one another. Moreover, anti-seizure medication has been associated with cognitive dysfunction, but evidence of these effects are mixed in part because of heterogeneities within the paediatric epilepsy population (53,54). To better understand the possible effects of epilepsy and its treatments on other aspects of *CASK*-related disorder, neurological data adopting standardised International League Against Epilepsy classification (55) and quantification of epilepsy severity and medication use plus longitudinal assessment of cognitive functions are required.

In contrast, we did not find descriptive differences in global adaptive ability between individuals with MICPCH and microcephaly only. This may suggest that cortical abnormalities, rather than hindbrain abnormalities, contribute to developmental delay and cognition in *CASK*-related disorder. This is inconsistent with the previous presumption that individuals with MICPCH will be more severely affected (12,20). However, we observed poorer average motor abilities in CYPs with MICPCH than those with microcephaly only, confirming contribution of cerebellar abnormalities to motor development. These findings are observational and MRI data was limited (only available for 50% of the BINGO *CASK*-related disorder sample). Future studies with comparable MRI data are needed to statistically interrogate the relationships between neuroanatomical abnormality and neurodevelopmental dimensions in *CASK*-related disorder.

## Conclusions

In conclusion, we provide an up-to-date description of the characteristics associated with *CASK*-related disorder, which includes a diverse range of clinical features, developmental abilities and social-emotional-behavioural characteristics. We suggest that the group-wise distributions of ID severity, neuroanatomical features and ophthalmological abnormalities are expanding with broader ascertainment of individuals with *CASK* variants. The relationships between structural brain development, epilepsy and developmental trajectories warrant further investigation in this group.

## Data Availability

Anonymised data may be made available to other researchers from the corresponding author on reasonable request, and on condition of signing a Code of Conduct guaranteeing that the data will be kept confidential and securely.

## Abbreviations

AAC: Augmentative and Alternative Communication
ASD: Autism Spectrum Disorder
B-H FDR: Benjamini-Hochberg False Discovery Rate
BINGO: Brain and Behaviour in Neurodevelopmental disorder of Genetic Origin
CASK: Calcium/calmodium-dependent serine protein kinase
CNV: Copy Number Variant
CYP: Children and Young People
CVI: Cerebral Visual Impairment
DBC2: Developmental Behaviour Checklist, Second Edition
FCVIQ: Flemish Cerebral Visual Impairment Questionnaire
ID: Intellectual Disability
MAGUK: Membrane-associated Guanylate Kinase
MHQ: Medical History Questionnaire
MICPCH: Microcephaly and pontine cerebellar hypoplasia
MRI: Magnetic Resonance Imaging
NGS: Next Generation Sequencing
PECS: Picture Exchange Communication System
RBQ: Repetitive Behaviour Questionnaire
SNV: Single Nucleotide Variant
SRS-2: Social Responsiveness Scale, Second Edition
VABS: Vineland Adaptive Behaviour Scale
XLID: X-linked Intellectual Disability

## Ethics declarations

### Ethics approval and consent to participate

Study protocol was approved by the Cambridge Central Research Ethics Committee (IRAS reference: 11/EE/0330, ‘Phenotypes in Intellectual Disability’), and written informed consent was obtained from each participant’s parent/caregiver prior to inclusion.

### Consent for publication

Not applicable

## Acknowledgements

We thank all of the children and young people with *CASK*-related disorder and their families for their participation in the BINGO project. We thank the charities who supported our research and recruitment of participants into the BINGO project, in particular CASK Research Foundation, Angelina CASK Neurological Research Foundation, Association Enfants CASK France, Unique, Cambridge Rare Disease Network, and Genetic Alliance UK.

## Author Information

### Authors and Affiliations

MRC Cognition and Brain Sciences Unit, University of Cambridge, 15 Chaucer Road, Cambridge, Cambridgeshire CB2 7EF

Jessica Martin, Alkistis Mavrogalou-Foti, Josefine Eck, Kate Baker

CASK Research Foundation, 33 Finchdean Road, Rowlands Castle, Hampshire PO9 6DA Laura Hattersley

Department of Medical Genetics, University of Cambridge, Addenbrooke’s Treatment Centre, Cambridge Biomedical Campus, Cambridge, Cambridgeshire CB2 0QQ

Kate Baker

Department of Pathology, University of Cambridge, Tennis Court Road, Cambridge, Cambridgeshire CB2 1QP

Kate Baker

### Contributions

JM and KB designed and conceptualised the study, with assistance of LH. JM led data collection, supported by AMF and JE. Data analysis and the literature review were conducted by JM and AMF. JM wrote the first draft of the manuscript. AMF wrote parts of the manuscript. KB provided supervision, and reviewed and edited the manuscript. All authors read and approved the final manuscript.

