## Supplementary Material 1 for "The neurodevelopmental spectrum of *CASK*-related disorder"

### **Supplementary Table 1.** Neurodevelopmental Questionnaire Measures.

| Questionnaire | Description | Scoring Procedure |
| --- | --- | --- |
| The Vineland Adaptive Behaviour Scales (VABS) (Third Edition, Parent/Caregiver Form, Vineland-3; 1) | The VABS is a standardised assessment of global adaptive ability, comprised of three subdomains (communication, daily living skills, and socialisation) and a separate measure of motor skills. | Raw VABS composite and subdomain scores were scaled according to published norms. For ease of interpretation, scores on the VABS composite were reversed, such that higher scores represent greater difficulties across measures. The VABS composite was categorised as ‘normal’, ‘mild’, ‘moderate’, ‘severe’ or ‘profound’, and was used here as a proxy measure of severity of developmental delay or intellectual disability (DD/ID). |
| Developmental Behaviour Checklist (DBC2; 2) | The DBC2 is a standardised measure of emotional-behavioural difficulties in individuals with ID. | T-scores for the total behavioural problems score (TBPS) and all 5 treatment subscales (disruptive/antisocial behaviour, self-absorbed behaviour, social relating, communication disturbance, and anxiety) were generated using the Australian norms. T-scores were not stratified for severity of ID due to missing VABS composite scores. |
| Social Responsiveness Scale (SRS-2; 3) | The SRS-2 is a screening tool designed to assess autism characteristics in the general population, and is comprised of 5 subscales (social awareness, social cognition, social communication, social motivation, and restricted interests and repetitive behaviour). | SRS-2 raw scores were converted to age- and sex-appropriate T-scores based on published norms. |
| Repetitive Behaviour Questionnaire (RBQ; 4) | The RBQ is a measure of repetitive behaviour frequency across 5 subscales (stereotyped behaviour, compulsive behaviour, insistence on sameness, restricted preferences, and repetitive use of language). | Scores on the RBQ were calculated using the verbal scoring approach and missing items were scored as 1 (never). |
| Flemish Cerebral Visual Impairment Questionnaire (FCVIQ; 5) | The FCVIQ is a measure of behaviours associated with CVI and its impact on everyday functioning. | The analysis here focused on total scores. For descriptive purposes, we generated the Sum Score described by Ortibus et al. (5). Using this score, an individual is considered at risk of CVI if they display a minimum of one behaviour in at least four of the FCVIQ domains. We also generated scores for the five factors identified by Ben Itzhak et al. (6) (object and face processing impairments, visual (dis)interest, clutter and distance viewing impairments, moving in space impairments, and anxiety-related behaviours), which we scored according to the method described by Crotti et al. (7). Higher scores on all FCVIQ measures reflect greater impairment. |
| Challenging Behaviour Questionnaire (CBQ; 8) | The CBQ is a measure of the presence and severity of behaviours that challenge (BtC), including self-injury, physical aggression, property destruction**,** and stereotypic behaviour. | The proportion of participants displaying each category of challenging behaviour was generated from parent-reported data indicating the presence of each category of behaviour in the past month on a yes/no basis. Severity scores for each category of challenging behaviour were calculated out of 14 based on the duration, frequency, severity of behaviour. Higher scores represent greater severity. This scoring approach is consistent with Bissell et al. (9). |
| Short Sensory Profile – 2^nd^ Edition (SSP-2; 10) | The SSP-2 measures sensory-related behaviours in daily life across four patterns of sensory processing - Seeking/Seeker, Avoiding/Avoider, Sensitivity/Sensor, and Registration/Bystander. These patterns are theorised to arise from the relationships between an individual’s neurological threshold for sensory stimulation, and self-regulation strategies in Dunn’s Sensory Processing Model (11). The SSP-2 also includes a total sensory and behavioural score. | Raw l subscale scores were calculated and classified according published norms. These scores were classified as “much less than others”, “less than others”, “just like the majority of others”, “more than others”, and “much more than others”, based on a bell curve normed distribution of scores from a sample of children without disabilities. |

### **Supplementary Table 3.** Behaviours that challenge and sensory-related behaviour in the BINGO *CASK*-related disorder group.

|  | | N | | M | | SD | | Range |
| --- | --- | --- | --- | --- | --- | --- | --- | --- |
| ***CBQ*** | | | | | | | | |
| Self-injury; n (present %) | 28 | | 12 (42.86%) | |  | |  | |
| Self-injury | 12 | | 5.75 | | 2.01 | | 3-8 | |
| Physical aggression; n (present %) | 28 | | 13 (46.43%) | |  | |  | |
| Physical aggression | 12 | | 5.25 | | 1.36 | | 3-8 | |
| Property destruction; n (present %) | 28 | | 8 (28.57%) | |  | |  | |
| Property destruction severity | 7 | | 5.71 | | 1.11 | | 5-8 | |
| Stereotyped behaviour; n (present %) | 28 | | 14 (50.00%) | |  | |  | |
| Stereotyped behaviour severity | 14 | | 4.86 | | 1.79 | | 2-8 | |
| ***SSP-2*** | | | | | | | | |
| Seeking | 26 | | 17.19 | | 6.07 | | 5-33 | |
| Avoiding | 26 | | 21.69 | | 10.00 | | 7-40 | |
| Sensitivity | 26 | | 29.88 | | 5.78 | | 16-41 | |
| Registration | 26 | | 19.92 | | 6.70 | | 3-30 | |
| Sensory | 26 | | 35.04 | | 9.50 | | 11-56 | |
| Behavioural | 26 | | 53.65 | | 15.5 | | 20-76 | |

### **Supplementary Table 4.** Classification of sensory-related behaviour on the SSP-2 in the BINGO *CASK*-related disorder group.

| SSP-2 Measure | SSP-2 Classification | | | | |
| --- | --- | --- | --- | --- | --- |
|  | Much Less Than Others | Less Than Others | Just Like the Majority of Others | More Than Others | Much More Than Others |
| Seeking | 0 (0.00%) | 1 (3.85%) | 13 (50.00%) | 11 (42.31%) | 1 (3.85%) |
| Avoiding | 0 (0.00%) | 5 (19.23%) | 8 (30.77%) | 7 (26.92%) | 6 (23.08%) |
| Sensitivity | 0 (0.00%) | 0 (0.00%) | 3 (11.54%) | 13 (50.00%) | 10 (38.46%) |
| Registration | 0 (0.00%) | 1 (3.85%) | 6 (23.08%) | 4 (15.38%) | 15 (57.69%) |
| Sensory | 0 (0.00%) | 1 (3.85%) | 7 (26.92%) | 12 (46.15%) | 6 (23.08%) |
| Behavioural | 0 (0.00%) | 0 (0.00%) | 8 (30.77%) | 7 (26.92%) | 11 (42.31%) |

### **Supplemental Table 5.** Correlations in the BINGO *CASK*-related disorder group.

|  | Age | Sex | VABS Composite | DBC2 Total | SRS-2 Total | RBQ Total | CVI Total |
| --- | --- | --- | --- | --- | --- | --- | --- |
| Sex | τ = -0.19, *p* = 0.22, *p.adj* = 0.39 |  |  |  |  |  |  |
| VABS Composite | *r_s_* = 0.78, ***p* < .01**, ***p.adj* < .01** | *r_pb_* = -0.21, *p* = 0.29, *p.adj* = 0.48 |  |  |  |  |  |
| DBC2 Total | τ = -0.03, *p* = 0.82, *p.adj* = 0.84 | *r_pb_* = -0.07, *p* = 0.70, *p.adj* = 0.77 | *r* = -0.33, *p* = 0.09, *p.adj* = 0.23 |  |  |  |  |
| SRS-2 Total | τ = 0.20, *p* = 0.14, *p.adj* = 0.31 | *r_pb_* = -0.14, *p* = 0.49, *p.adj* = 0.62 | *r* = 0.14, *p* = 0.49, *p.adj* = 0.62 | *r* = 0.36, *p* = 0.06, *p.adj* = 0.22 |  |  |  |
| RBQ Total | τ = 0.09, *p* = 0.49, *p.adj* = 0.62 | τ = -0.32, ***p* = 0.04**, *p.adj* = 0.22 | τ = -0.09, *p* = 0.51, *p.adj* = 0.62 | τ = 0.55, ***p* < .01**, ***p.adj* < .01** | τ = 0.24, *p* = 0.09, *p.adj* = 0.23 |  |  |
| CVI Total | *r_s_* = -0.16, p = 0.51, p.adj = 0.62 | *r_pb_* = 0.24, *p* = 0.31, *p.adj* = 0.48 | *r* = 0.05, *p* = 0.84, *p.adj* = 0.84 | *r* = 0.33, *p* = 0.16, *p.adj* = 0.31 | *r* = 0.50, ***p* = 0.03**, *p.adj* = 0.21 | τ = -0.21, *p* = 0.20, *p.adj* = 0.38 |  |
| Epilepsy | τ = 0.29, *p* = 0.07, *p.adj* = 0.22 | *p* = 0.62, *p.adj* = 0.73^a^ | *r_pb_* = 0.55, ***p* < .01**, ***p.adj* = 0.03** | *r_pb_* = -0.34, *p* = 0.07, *p.adj* = 0.22 | *r_pb_* = 0.08, *p* = 0.71, *p.adj* = 0.77 | *r_pb_* = -0.34, *p* = 0.07, *p.adj* = 0.22 | *r_pb_* = 0.33, *p* = 0.16, *p.adj* = 0.31 |

τ Kendall; r_s_ Spearman; r_pb_ point-biseral ^a^Fisher’s exact test

### **Supplemental Table 6a.** Impact of Epilepsy on Adaptive Ability: Results of Regression Assumption Tests.

| Assumption | Test | Result |
| --- | --- | --- |
| Independence of Errors | Durbin-Watson | D-W = 2.31, *p* = .43 |
| Homoscedasticity | Breusch-Pagan | χ²(2) = 1.43, p = .23 |
| Normality of Residuals | Shaprio-Wilk | W = .98, p = .79 |

### **Supplemental Table 6b.** Impact of Epilepsy on Adaptive Ability: Regression Analysis Results.

| Variable | B | SE | t | p | 95% CI |
| --- | --- | --- | --- | --- | --- |
| Constant | 89.63 | 3.57 | 25.11 | 2e-16 | [82.27, 96.98] |
| Epilepsy: Yes | 18.28 | 5.59 | 3.27 | 0.00313 | [6.77, 29.80] |
| R2 | 0.30 |  |  |  |  |
| Adj.R2 | 0.27 |  |  |  |  |

### **Supplemental Table 7a.** Impact of Epilepsy and Age on Adaptive Ability: Results of Regression Assumption Tests.

| Assumption | Test | Result |
| --- | --- | --- |
| Independence of Errors | Durbin-Watson | D-W = 1.95, *p* = .79 |
| Homoscedasticity | Breusch-Pagan | χ²(2) = 5.05, p = .08 |
| Normality of Residuals | Shaprio-Wilk | W = .96, p = .38 |
| Multicolliniarity | Variance Inflation Factors | All VIFs < 10 |

### **Supplemental Table 7b.** Impact of Epilepsy and Age on Adaptive Ability: Regression Analysis Results.

| Variable | B | SE | t | p | 95% CI |
| --- | --- | --- | --- | --- | --- |
| Constant | 77.40 | 4.18 | 18.50 | 1.04e-15 | [68.76, 86.03] |
| Epilepsy: Yes | 11.00 | 4.80 | 2.81 | 0.032 | [1.04, 20.87] |
| Age | 1.56 | 0.39 | 3.97 | 0.00057 | [0.75, 2.38] |
| R2 | 0.58 |  |  |  |  |
| Adj.R2 | 0.54 |  |  |  |  |

### **Supplemental Table 8.** Summary of characteristics by brain abnormality information for the BINGO *CASK*-related disorder group.

|  | MICPCH (n = 13)^a^ | | | Microcephaly (n = 9)^b^ | | | Normal (n = 2)^c^ | | | NA (n = 6)^d^ | | |
| --- | --- | --- | --- | --- | --- | --- | --- | --- | --- | --- | --- | --- |
|  | *M* | sd | Range | *M* | sd | Range | *M* | sd | Range | *M* | sd | Range |
| Age / years | 9.76 | 6.23 | 3.04-23.88 | 9.44 | 4.57 | 4.10-15.80 | 12.83 | 0.10 | 12.76-12.90 | 9.41 | 8.40 | 3.10-25.86 |
| Gender; n (% female) | 13 (100.00) | | | 7 (77.78) | | | 1 (50.00) | | | 5 (83.33) | | |
| VABS Composite^e^ | 40.15 | 15.25 | 20-68 | 43.62 | 14.24 | 22-61 | 59.00 | 21.21 | 44-74 | 50.4 | 21.73 | 20-75 |
| VABS Motor^e^ | 30.54 | 15.72 | 20-68 | 46.88 | 8.41 | 33-62 | 64.5 | 23.33 | 48-81 | 44.6 | 19.06 | 20-71 |
| CVI Total^f^ | 19.50 | 7.11 | 4-33 | 15.80 | 6.02 | 10-24 | 15.00 | 1.41 | 14-16 | 12.33 | 10.07 | 3-23 |
| Epilepsy; n (% present) | 7 (53.85) | | | 3 (33.33) | | | 0 (0.00) | | | 1 (20.00) | | |

^a^ Included participants who have reported MICPCH (note: one participant had CH/PH only, and one participant had MIC and CH only).

^b^ Included participants who have reported microcephaly (note: three participants have reported microcephaly and no available MRI data, one participant has reported microcephaly and unclear MRI data; four participants have reported microcephaly and normal MRI, and one participant has microcephaly and agenesis of corpus callosum).

^c^ Included participants whom have no reported microcephaly, or structural brain abnormalities, , either via MRI report, genetic report, or parent report.

^d^ Included participants for whom no brain abnormality data is available, either via MRI report, genetic report, or parent report.

^e^ Variable n - microcephaly n = 8, NA n = 5.

^f^ Variable n - MICPCH n = 10, Microcephaly n = 5, NA n = 3.
